# Improved NAAT assay for the diagnosis of onchocerciasis and its use for detection of circulating cell free DNA

**DOI:** 10.1101/2023.09.13.23295358

**Authors:** Sasisekhar Bennuru, Frimpong Kodua, Eric Dahlstrom, Thomas B Nutman

## Abstract

Mass drug administration (MDA) programs aimed at control and elimination of onchocerciasis relies on annual or semi-annual distribution of Ivermectin (IVM) that target the microfialarial (mf) stage of the parasite *Onchocerca volvulus*. The co-endemicity of onchocerciasis with other filarial species often leads to interruptions in the control programs. A much-needed tool for the elimination efforts are sensitive diagnostic assays that can help differentiate from other co-endemic filarial infections or xenomonitoring approaches. qPCR assays targeting highly repeated elements in the genomes have allowed for increased sensitivity of detection that are pathogen-specific. Utilizing NGS data, qPCR assays were designed to target 15 highly repeated targets from *O. volvulus* and 11 from *O. ochengi*. The two most sensitive and specific repeats Ov15R and Ov16R from *O. volvulus* and OoR1 and OoR5 from *O. ochengi*, were selected for further testing.

The analytical sensitivity of both Ov15R and Ov16R were similar with limits of detection (LOD) at 1 fg of *O. volvulus* genomic DNA (gDNA), and 100% specificity and ∼32-fold sensitive than O-150 qPCR. Similarly, OoR1 and OoR5 had LOD of 100 fg of *O. ochengi* gDNA. Using DNA obtained previously from skin snips, Ov16R identified 17 additional samples as positive for infections when compared to O-150.

Further, to eliminate the need for time-dependent sample acquisition, we evaluated the efficiency of plasma-derived circulating cell-free DNA (ccfDNA) to detect *O. volvulus* infections and as a potential modality for testing cure of infection. Plasma-derived ccfDNA failed to amplify Ov16R. In contrast using a small set of urine-derived ccfDNA samples from Ov-infected individuals had varying levels of detectability as early as 12-24 hours post-treatment. To enable processing of larger volumes of urine for better sensitivity, a modified chitosan-based filter technique was developed as proof-of-concept that efficiently captured gDNA from 1-15ml of urine.

Interestingly, Ov15R, Ov16R and O-150 map to the same contigs of *O. volvulus* genome. This prompted the redesign of O-150 qPCR that resulted in O-150-New qPCR assay that performs on par with Ov15R/Ov16R, thus offering more sensitive and specific diagnosis of *O. volvulus* that can easily be configured to field-friendly formats such as LAMP or RPA.

## Introduction

*Onchocerca volvulus* is a filarial parasite that causes onchocerciasis or more often known as ‘river blindness’ and is transmitted by the bites of *Simulium* blackfly species primarily endemic to sub-Saharan Africa. Control efforts for elimination of the disease as a major public health concern relies on the mass drug administration (MDA) programs with annual or semi-annual distribution of Ivermectin (IVM). Only the microfilarial (mf) stages of the parasite are effectively targeted using IVM that help to reduce disease burden and block transmission to the blackfly vectors (*Simulium* sp). Based on the microfilarial prevalence, endemic regions can be broadly categorized into hypoendemic (<30-35%), mesoendemic (30-35% to 60%) and hyperendemic (≥ 60%).

A much-needed tool for the elimination efforts is a specific and sensitive diagnostic assay that can verify if the levels of transmission of the disease are low enough to warrant the cessation of MDA or in regions of post-MDA surveillance for monitoring resurgence of infection. The use-case scenario for the assay relies on the assay(s) to have high sensitivity and specificity, to detect very low-levels of mf densities while being able to discriminate between closely related co-endemic major filarial parasites that infect humans such as *Loa loa*, *Wuchereria bancrofti, Mansonella perstans* and the bovine filarial parasite *Onchocerca ochengi* that is also transmitted by the same vector species. Another major concern is the presence of *Loa loa* microfilariae in the skin-snips of infected individuals in co-endemic areas [1] that often lead to false interpretations.

Historically, the identification of microfilariae in the skin snips by microscopy was the ‘gold standard’ for diagnosis of infection. Though skin snip microscopy is 100% specificity it is plagued with low sensitivity (estimated at 20%) esp. in areas with low mf densities[2].

The guidelines from World Health organization (WHO) for stopping MDA and verifying elimination of onchocerciasis as a major public health concern recommends serological evaluation for Ov16 by ELISA or rapid diagnostic test (RDT) [3], where the seropositivity to Ov16 in children younger than 10 years should be <0.1% and positivity of pool screen PCR of at least 6000 black fly heads to be <0.05%, the diagnostic performance thresholds of which depend on multiple factors [3, 4]. The Ov-16 antibody test determines the presence of IgG4 antibodies to Ov-16 as a marker of exposure to the parasite [5–8]. Entomological screening for the presence of infective L3 larvae in the vector population relies on the PCR based amplification of O-150 repeat family region of parasite DNA [9–11]. Though there is some conservation of the O-150 tandem repeats sequences across the *Onchocerca* spp., the tandem repeats not only vary in the number but also not always identical repeats. In addition to the detection of O-150, assays based on real-time qPCR and loop mediated isothermal amplification (LAMP) have been developed to detect *O. volvulus* DNA in the skin [12]. Skin snip evaluation by PCR could be used to also differentiate active infection from exposure to the parasite where the prevalence of Ov-16 seropositivity is > 0.1% [3]. However, skin snips being more invasive in nature are becoming increasingly unpopular and often refused by endemic communities [13]. Recent studies explored the detectability of circulating filarial DNA for *B. malayi* [14], *L. loa* [15] or *Onchocerca* spp [16]. Though parasite-derived miRNAs were detected in the plasma of *O. volvulus* infected individuals, their detection as a biomarker has been challenging and were determined to be insufficient tools or as biomarkers of treatment efficacy for *O. volvulus* [16, 17]. Detecting circulating cell-free DNA in body fluids (serum/plasma or urine) provides a promising alternative where sample collection is more convenient or less invasive.

In this study, we explored the utility of repeatexplorer pipeline that has previously been shown to identify targets that are not only more sensitive in detecting infections [15, 18–21] and specific, but also enable the monitoring of treatment efficacy [15].

## Methods

### Samples

All archived serum/plasma or urine samples were obtained as part of National Institute of Allergy and Infectious Diseases’ Institutional Review Board-approved protocols for filaria-infected patients (NCT00001230) and healthy donors (NCT00090662), that were from previous studies [22, 23] and stored at −80C. As part of routine clinical care, informed written consent was obtained from all subjects. Archived DNA samples from skin snips were from previous studies [1], that received ethical clearance from the Cameroon National Ethics Committee for Research in Human Health (No. 2013/11/370/L/CNERSH/SP), and an administrative authorization was granted by the Ministry of Public Health (D30-571/L/MINSANTE/SG/DROS/CRSPE/BBM). All eligible volunteers signed an informed consent form for their participation in the study, including for the further use of their samples.

### Target identification and Assay Design

Sequence read archives (SRA) from BioProjects PRJEB513, PRJEB17785 and PRJNA289926 for *O. volvulus* and BioProject PRJEB23566 for *O. ochengi* were processed to trim, clean and obtain paired reads. The RepeatExplorer [24, 25] pipeline was used to analyze the paired reads and generate highly repeated contigs (numbered with a prefix of “Ov” and suffix “R”). The most promising contigs for *O. volvulus* (n=15) and *O. ochengi* (n=11) with >1000x coverage, minimal length of 100 base pairs and minimal homology with other helminths and human were selected (**Table S1**). O-150 qPCR was also redesigned to improve the assay. PCR assays were designed for these selected sequences using the PrimerQuest Tool (Integrated DNA Technologies, IDT), and custom oligos synthesized for screening with real-time qPCR assays.

### ccfDNA extraction and real-time PCR

Circulating cell-free DNA was extracted from 100, 250, 500 or 1000 μl of plasma using the MinElute ccfDNA kit or its automated version Viral/Pathogen DSP kit for Qiasymphony. The samples were eluted in a final volume of 20 µl for MinElute and 60 µl on the Qiasymphony.

DNA extractions from 1 ml of urine were as mentioned above, while larger volumes were performed using a chitosan-modified filter paper technique [26, 27] with slight modifications. Briefly, Fusion 5 filter paper (Whatman) were washed in 30 ml 0.2 M NaOH for 5 minutes, washed with 50 ml deionized water (3x) and dried under vacuum at 45°C. The filters were soaked in chitosan (0.25%) dissolved in 0.1 M acetic acid for 16 hours at room temperature on a rotary shaker. The membranes were washed (3x) in deionized water and dried under vacuum at 45°C. 4-mm diameter discs of chitosan-functionalized membranes and Whatman GB003 cellulose paper were made using biopsy punch. A 2 mm-diameter opening was made in a female Luer cap (Cole Parmer, Vernon Hills, IL) using a biopsy punch. The 4-mm GB003 cellulose membrane disc (as backing support membrane) followed by the chitosan-functionalized disc were positioned in the luer cap. Urine samples mixed 1:1 with 50 mM MES (pH 5.0) were passed through the luer caps using 20 ml syringes at a flow rate of 0.5 ml per minute using the PHD ULTRA (Harvard Apparatus). The DNA trapped on the chitosan-functionalized membrane was retrieved by boiling the disc in 50 μl of deionized water.

qPCR was performed in quadruplicates using an ABI 7900 sequence detection system using Taqman fast chemistry reagents (Applied Biosystems,Carlsbad, CA, USA) and primer/probe sets described in **Table 1**. Amplification conditions were 20 seconds at 95°C followed by 40 cycles of 1 second at 95°C and 20 seconds at 60°C. An internal plasmid control [28] was spiked into all samples prior to ccfDNA extraction to exclude false negatives due to qPCR inhibition. For a sample to be considered as positive at least three of the 4 replicates needed to positive. An internal plasmid control [28] was spiked into all samples prior to ccfDNA extraction to exclude false negatives due to qPCR inhibition.

### Statistical analyses

All statistical analyses were performed using GraphPad Prism v8. Unpaired samples were compared using Mann-Whitney test and paired analyses were performed with Wilcoxon matched-pairs signed rank test.

## Results

### O. volvulus and O. ochengi target identification

The repeatexplorer pipeline [24, 25] resulted in the identification of several highly repetitive contigs in the genomes of *O. volvulus* and *O. ochengi* of which, thirteen of the most highly represented ones for *O. volvulus* and eleven of the *O. ochengi* contigs were selected to be screened by qPCR (**Figure 1**). Preliminary screening with 10-fold dilutions of 1ng/ul of *O. volvulus* genomic DNA (Ov gDNA) indicated that Ov15R, Ov16R, Ov21R, Ov31R, Ov36R and Ov54R had better sensitivity than the other repeat contigs (**Figure 1A**). Moreover, these six assays had better sensitivity (lower Ct values) compared to the current gold standard O-150 qPCR (**Figure 1B**), of which Ov15R and Ov16R were chosen for being the most sensitive assays. Ov15R and Ov16R assays were similar in their limits of detection (1fg of gDNA) while maintaining specificity with gDNA of *B. malayi, L. loa, W. bancrofti, M. perstans* and *O. ochengi* (**Figure 1C**).

**Figure 1.**
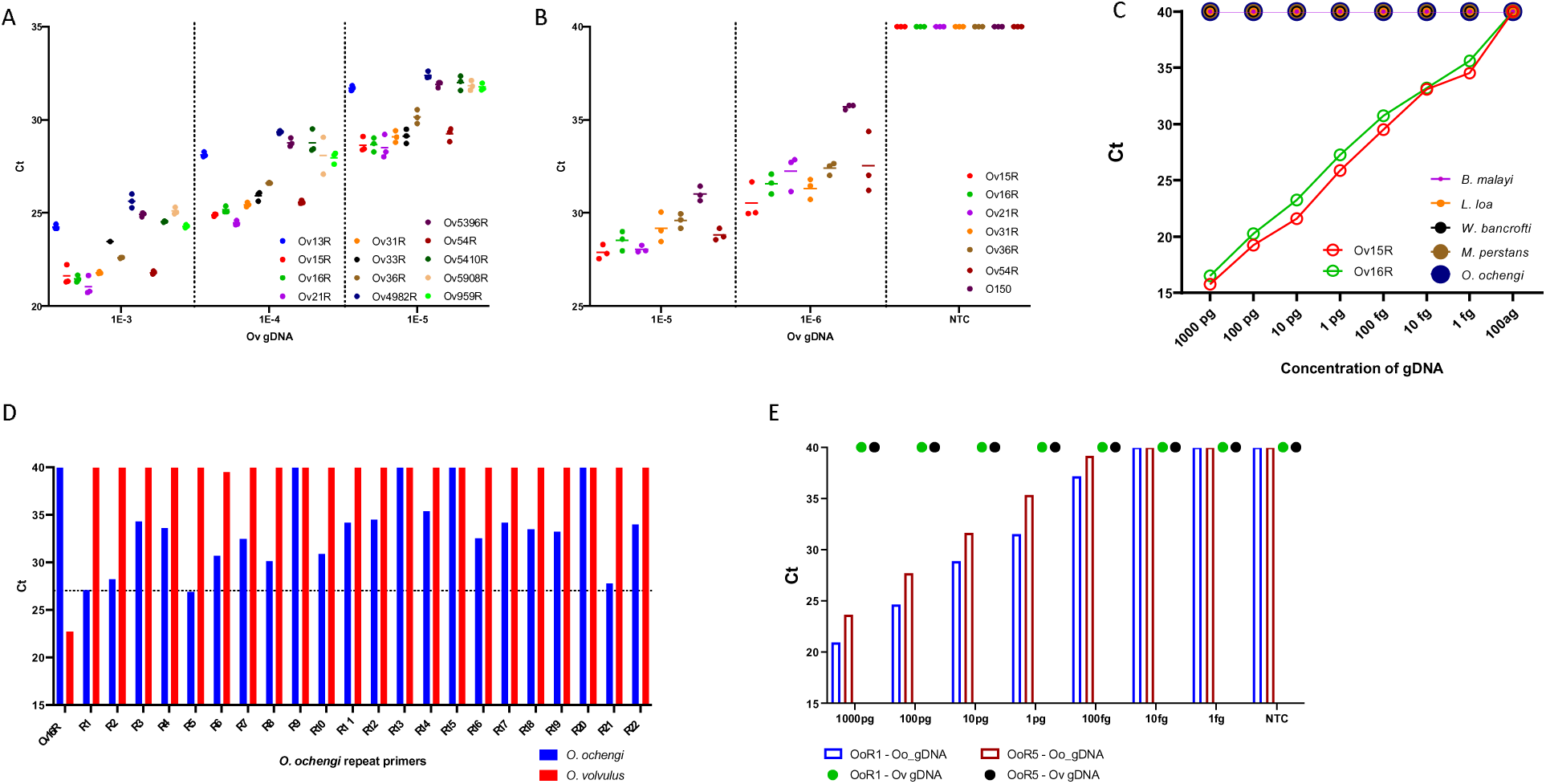
Identification of *Onchocerca volvulus* and *Onchocerca ochengi* repeat elements. (A) The graph depicts the screening of the identified repeat elements for limits of detection (plotted as Ct-values on the y-axis) by qPCR using dilutions of *O. volvulus* gDNA. (B) The graph represents the efficiency of the top five assays targeting repeated elements in comparison with O-150. (C) The graph depicts the specificity of Ov16R and Ov15R with serial dilutions of gDNA across major filarial parasites of humans. (D) The bar graph depicts the screening of the identified repeat elements for limits of detection (plotted as Ct-values on the y-axis) by qPCR using *O. ochengi* (blue) and *O. volvulus* (red) gDNA. (E) The bar graph shows the limits of detection (plotted as Ct-values on the y-axis) of *O. ochengi* repeat assays (OoR1 and OoR5) by qPCR using dilutions of *O. ochengi* (red and blue) and *O. volvulus* (green and black) gDNA. No amplifications were set to 40 cycles.

The eleven *O. ochengi* repeat contigs were screened with 2 assays designed for each contig (R1-R22), of which R1 and R5 (henceforth referred to as OoR1 and OoR5) were the most sensitive (lowest Ct value) when screened with 10 pg of *O. ochengi* gDNA (**Figure 1D**). Screening of OoR1 and OoR5 with gDNA standards indicated the LOD of 100 fg with *O. ochengi* gDNA while maintaining specificity with *O. volvulus* gDNA (**Figure 1E**).

### Relationship between O-150 and Ov15R/Ov16R

Because Ov15R and Ov16R had better sensitivity (lower Ct values) compared to O-150 qPCR assay (**Figure 2A**), combinations of Ov15R and/or Ov16R were tested with O-150 but did not yield any enhancements in the limits of detection. Sequence analyses suggested that the selected *O. volvulus* contigs might be related somehow to O-150 in that they all mapped to one end of OVOC_OO_000013, OVOC_OO_00548 or OVOC_OM1b of the *O. volvulus* genome. This prompted a re-designing of the O-150 assay, where all the new assays (O-150 N1-N5) performed much better than the original O-150 assay and comparable to Ov15R/Ov16R (**Figure 2B**). The O-150 N1 (henceforth called O-150New) was used for all further comparative analyses when needed. It is not clear as to what drives this new assay to be more sensitive but compared to the original oligos, the major observable change was the number of locations that O-150New reverse could bind in the O-150 repeat region (Acc. No. J04659.1) (**Figure S1**). Since Ov15R and Ov16R were essentially similar, all further analyses were done with Ov16R.

**Figure 2.**
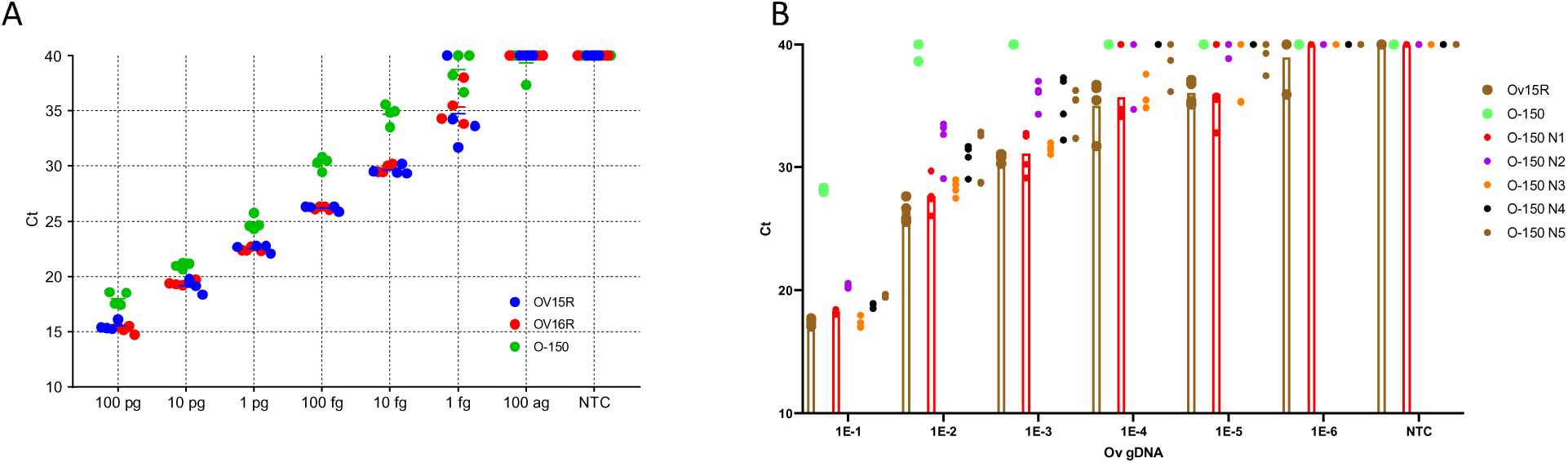
Development of new assays for *O. volvulus. (A)* The graph represents the efficiency of the Ov16R (red) and Ov15R (blue) in comparison with O-150 (green) across different concentrations of *O. volvulus* gDNA. (B) Screening of re-designed O-150 new assays (O-150 N1-N5) for limits of detection (plotted as Ct-values on the y-axis) in comparison with Ov15R and current O-150 qPCR assay

### Efficiency of Ov16R with skin snip samples

The efficacy of Ov16R was evaluated with archived DNA extracted from skin snip samples from two previous studies: 1) individuals with nodding syndrome that were previously screened with O-150 qPCR and 2) from individuals living in an area co-endemic for loiasis. As shown in **Figure 3A**, of the 95 samples with nodding syndrome, 42 (44%) were positive by O-150. Ov16R with slightly better (lower) Ct values (**Figure 3B**) was able to pick up 17 additional samples as positive for a total of 59 (62%) positives (**Figure 1C**). Likewise, DNA extracted from skin snip samples from an area co-endemic for loiasis with microfilariae in the skin snips that were previously diagnosed for the vast majority as microfilariae of *Loa loa* and not *Onchocerca volvulus*, were tested with Ov16R. As shown in **Figure 3D**, despite a modest gain in Ct values compared to O-150, there were no additional positives that were detectable with Ov16R. As expected, the O150 results correlated similarly with O-150-New and Ov16R Ct values, with O-150-New performing similarly to Ov16R in comparison to the O-150 (**Figure 3E-G**).

**Figure 3.**
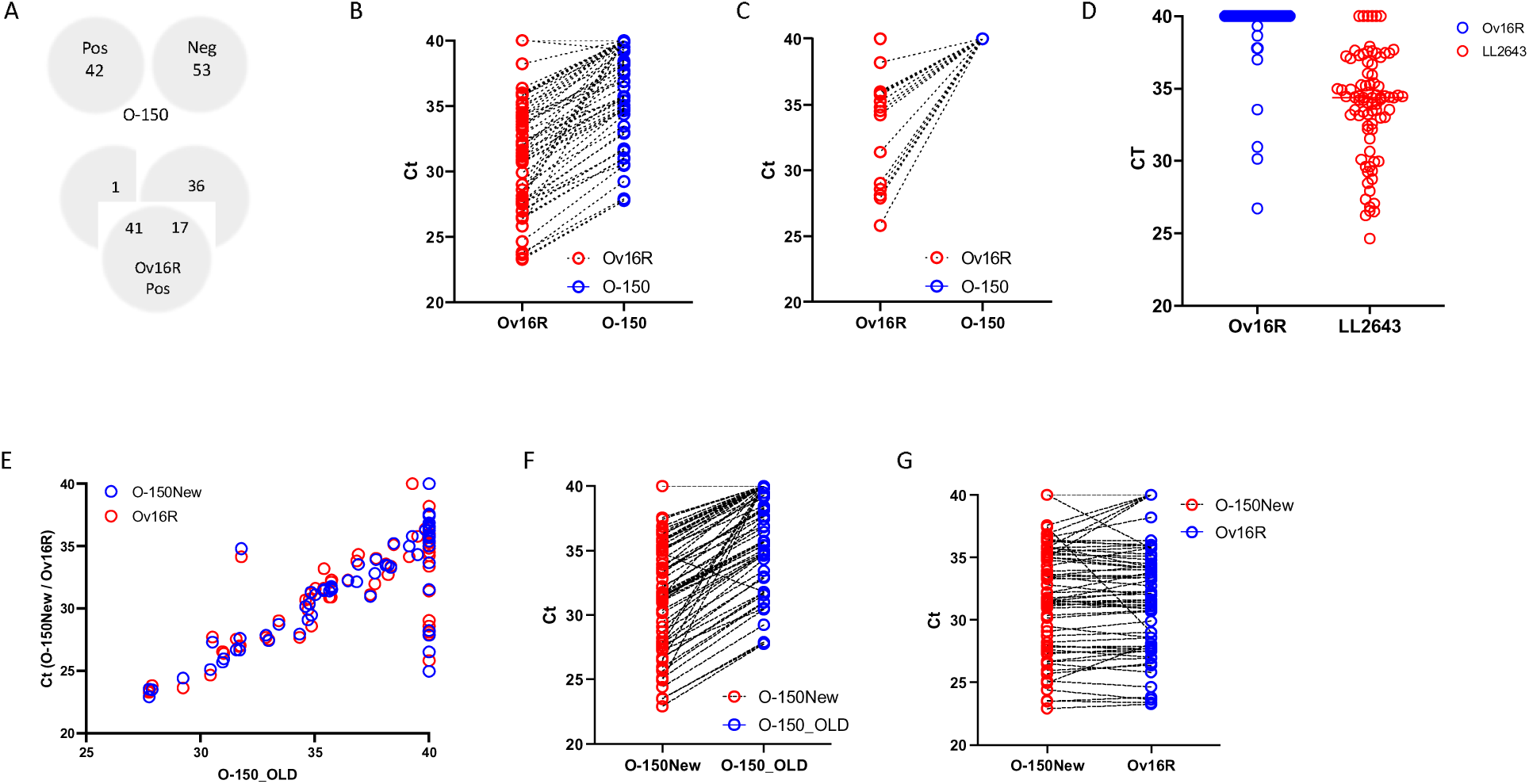
Efficiency of Ov16R and O-150New *(A)* The VENN diagram denotes the O-150 qPCR status of skin snips (top panel), with Ov16R detecting 17 additional samples missed by O-150. (B/C) The graph represents the paired Ct values obtained by Ov16R and O150, and the 17 additional samples being detected by Ov16R that were O-150 negative. (D) Ov16R sensitivity is improved (Ct values) compared to O-150 but does not detect any new additional samples from *Loa loa* co-infected area. (E) Graph depicts the correlation of the O150 assay with that of O-150New and Ov16R. (F&G) The graph shows the paired comparison of O150New with O-150 (O-150_OLD) or Ov16R.

### Plasma circulating cell-free DNA (ccfDNA)

To evaluate the efficacy of detecting Ov16R or O-150-New in ccfDNA derived from plasma of infected individuals, pooled plasma of infected individuals or pooled plasma of health blood bank individuals spiked with 100 pg of gDNA were used to extract ccfDNA. The ccfDNA extraction was carried out using plasma volumes of 100 µl, 250 µl, 500 µl and 1000 µl, using the MinElute ccfDNA kit (Qiagen) with elution volumes of 20 µl for all samples. As shown in **Figure 4A**, ccfDNA derived from any of the pooled samples (and volumes) from Ecuador or Guatemala failed to amplify Ov16R. As expected, the ccfDNA from pooled blood bank individuals were negative but the spiked samples were highly positive for Ov16R, suggesting that the lack of detectability is likely due to the absence of detectable levels of parasite-derived DNA in circulation.

**Figure 4.**
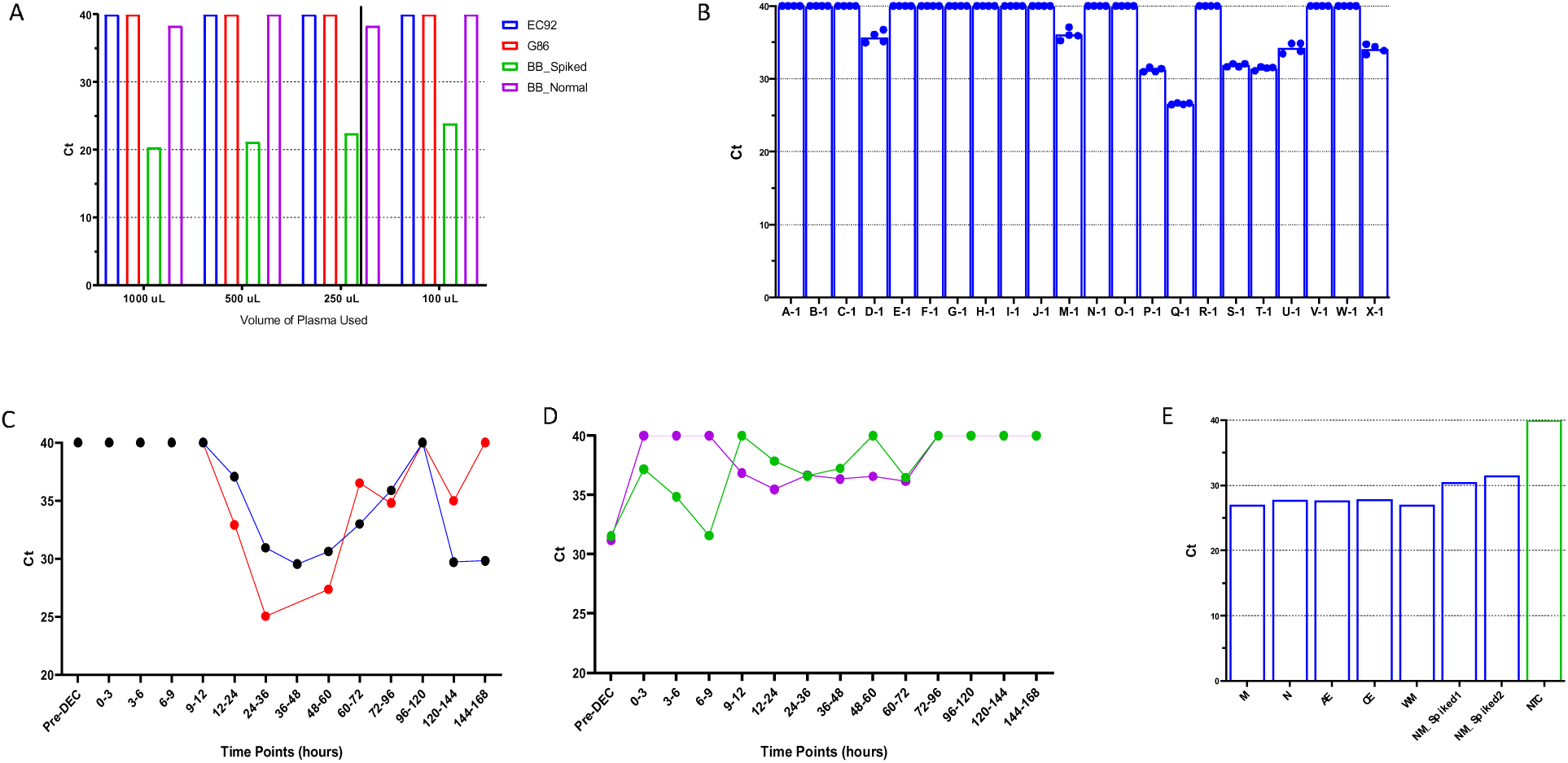
Efficacy of ccfDNA from plasma and urine. (A) the graph represents the Ov16R qPCR status (Ct values) of ccfDNA extracted from 1000 ul, 500 ul, 250 ul and 100 ul of pooled sera (n=10) from Ov-infected individuals from Guatemala (G86), Ecuador (EC92) or pooled plasma of healthy blood bank volunteers (n=10) with or without Ov-gDNA spiked. (B) the graph represents the Ov16R qPCR status (Ct value) with ccfDNA extracted from 1 ml of urine from Ov-infected individuals before treatment (C&D) the time course results of Ov16R qPCR with ccfDNA extracted from post-treatment samples (E) the graph represents the Ov16R status of 5 fresh urine samples from Cameroon.

### Urine circulating cell-free DNA (ccfDNA)

Urine as a source of detecting the presence of ccfDNA was tested using archived samples from previous studies (ref), that was aimed at understanding the immunological sequelae during Mazzotti reactions post treatment with ivermectin and followed up every 3-12 hours. ccfDNA was extracted from 1 ml of urine from 22 samples pre-treatment (18-infected and 4-controls). As shown in **Figure 4B**, 8/18 (44%) samples were positive for Ov16R at time 0. Since ivermectin targets only the microfilarial stages, and the death of microfilariae would result in parasite-derived DNA in circulation, we screened the time-course samples for 4 individuals that were still available (2 each for samples that were negative or positive at time 0). As expected, in the two samples that were negative pre-treatment, Ov16R could be detected as early as 12-24 hours post-treatment (**Figure 4C**), that eventually clears away around 3-4 days post-treatment. In contrast, the two samples that were positive for Ov16R pre-treatment had varying levels of detectability all through the time-course (**Figure 4D**).

To evaluate if there would be any difference in the sensitivity of ccfDNA in urine that has been frozen away for two decades compared to fresh urine, we obtained more recent/fresh urine samples from 5 individuals from Cameroon. Surprisingly, ccfDNA extracted from 1 ml of all five urine samples were positive for Ov16R (**Figure 4E**).

### Chitosan filter-based capture of ccfDNA

Though urine is not exactly a limiting factor in terms of sample volume, the processing of larger volumes with existing kits/reagents render it highly unusable. Hence to evaluate the feasibility of capturing/trapping the ccfDNA, we adapted the chitosan-modified filter paper technique (ref). Because of the non-availability of patient derived urine samples in higher volumes, pooled normal urine (1 or 15 ml) was spiked with various concentrations of *O. volvulus* gDNA and an internal control (IAC) and ccfDNA extracted onto chitosan filters (see methods). In parallel, 1 ml of urine samples with the spiked gDNA was also extracted on QiaSymphony. The filters were boiled in 30 ul of TE buffer (pH 8.0) to elute the DNA for qPCR detection. As shown in **Figure 5**, the efficiency of gDNA captured from 1 ml or 15 ml of urine and subsequent detectability of Ov16R were identical, indicating that it is possible to filter out larger volumes of urine, while capturing DNA.

**Figure 5.**
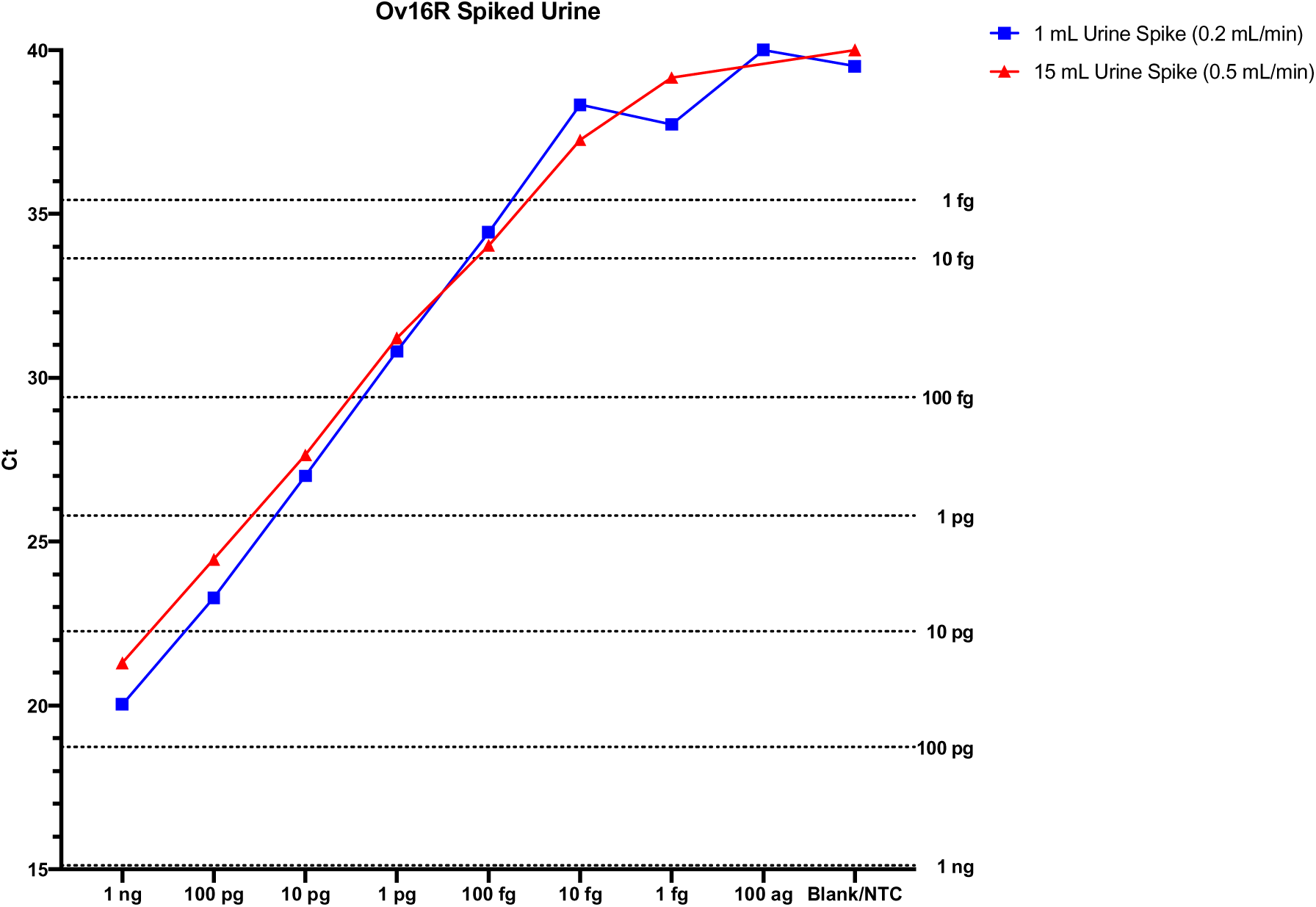
Efficacy of ccfDNA from chitosan-modified filter membrane. The graph shows the Ov16R qPCR (Ct values) of DNA extracted from 1 ml or 15 ml of normal urine spiked with varying concentrations of gDNA. The dotted lines denote the Ct values of respective concentrations of gDNA.

## Discussion

This study reports the identification of genomic repeats enriched in the genomes of *O. volvulus and O. ochengi.* Among those identified for *O. volvulus*, Ov16R was more sensitive than O-150 by qPCR. Based on informatic analyses, O-150 and Ov16R are related by way of their genomic location as they both map to one end of OVOC_OO_000013, OVOC_OO_00548 or OVOC_OM1b of the *O. volvulus* genome. The superiority of Ov16R to O-150 qPCR in terms of sensitivity was probably overcome using RepeatExplorer pipeline that utilizes NGS data to directly infer repeated elements that have been successfully utilized for developing highly sensitive diagnostic assays [18–20], instead of assembled contigs of the genome, where highly repetitive sequences in genome assemblies are often omitted as orphan DNA during the assembly process.

Diagnostic testing for onchocerciasis in regions co-endemic for *Loa* or *Mansonella* spp can be challenging as mf of *M. streptocerca* typically migrate through the skin [29], whereas mf from *M. perstans* and *L. loa* circulate in the vasculature [30]. Nevertheless, microscopy analyses of *L. loa* can be misdiagnosed as mf of onchocerca in nature especially in cases where the burden is high [1]. With the availability of better repeat targets for detecting *L. loa* [15], *M. perstans* [21], the combination of Ov16R or O150New would be a great tool to not only help in more accurate diagnosis but to also advance the global efforts for mapping and elimination of neglected tropical diseases in areas co-endemic for filarial infections. Another bottleneck in the molecular xenodiagnoses or xenomonitoring is the presence of other onchocerca species (*O. ochengi*, *O. guttarosa* etc) in the vectors. The use of OoR1 or OoR5 along with Ov16R or O150New would help differentiate *O. ochengi* from *O. volvulus*). Alternatively, the use of OvND5 (GenBank: AY462885.1 and FM206483.1) [31] that is slightly less sensitive but highly specific for *O. volvulus* or *O. ochengi* could be implemented as a second or confirmatory test to rule of *O. volvulus*.

Circulating cell-free DNA (ccfDNA) are residual fragments of DNA present in the peripheral circulation and have been used as good biomarkers in pregnancy, diabetes, cancer and solid-organ transplantation (reviewed in [32–34]). Since the circulating cell-free DNA in the body fluids (plasma and/or urine) are short-lived and likely to be excreted in urine, we evaluated the ability to detect Ov16R in plasma- and urine-derived ccfDNA of infected individuals to reflect the presence of active infection as the parasites release DNA (homeostatic and upon injury/death after treatment). However, we were unable to detect Ov16R reliably in ccfDNA derived from 250 ul of plasma. Increasing the initial of plasma to 2 ml did not alter the outcome significantly, suggesting that the inability to detect Ov16R in plasma-derived ccfDNA may partly be due to the niche of *O. volvulus* (skin-dwelling) in contrast to the blood vasculature dwelling filarial parasites (*W. bancrofti*, *B. malayi* and *L. loa*). It is interesting that the novel repeat regions that were identified earlier were also located in contigs that shared the O-150 repeat [16], but were no better at detecting circulating parasite-derived DNA.

The kinetics of detecting Ov16R in urine after treatment with DEC and their disappearance after 72 hours correlates well with the rapid death of the microfilariae and subsequent clearance from circulation with the immunohistological and microscopical observations that the mf disappear within 3-days of microfilaricidal treatment [35, 36].

Nevertheless, in contrast to *L. loa* infections [15], urine proved to be a better source for parasite-derived ccfDNA. While the lack of parasite derived ccfDNA in plasma could be attributed to the niche of *O. volvulus* (in contrast to vascular dwelling filaria such as *W. bancrofti* or *L. loa*), it is not clear as to why Ov16R was more readily detectable in the limited number of more recent/fresh urine samples compared to archived frozen urine samples. Previous studies demonstrated that the ccfDNA is comprised of highly fragmented double stranded DNA around 150bp in length, that relates to about 9,000-17,000 genome equivalents of human diploid genome in 1 ml of blood [37, 38]. Though there is no established method of characterizing the nature of parasite-derived ccfDNA in circulation, they are comparatively less abundant when compared to the host-derived ccfDNA. The rapid turnover and estimated renal clearance of ∼30 minutes is an important feature of ccfDNA that can be exploited for active cases of infection. The ccfDNA content is also known to vary as a function of collection time, duration interval and storage conditions [39]. Prospectively, analyzing larger volumes of urine samples using the chitosan filter-based method needs to be evaluated for the efficiency of DNA capture. Moreover, once optimized, this would also facilitate easy storage of the filters compared to storing larger volumes in freezers.

## Supporting information

Primers and probes

## Data Availability

All data produced in the present work are contained in the manuscript

## Acknowledgements

This work was supported in part by the Division of Intramural Research (DIR) of the National Institute of Allergy and Infectious Diseases, NIH.

We thank Dr. Sara Lustigman from New York Blood Center for fresh urine samples from Cameroon.

All data produced in the present work are contained in the manuscript

## REFERENCES

1. Nana-Djeunga HC, Fossuo-Thotchum F, Pion SD, Chesnais CB, Kubofcik J, Mackenzie CD, et al. Loa loa Microfilariae in Skin Snips: Consequences for Onchocerciasis Monitoring and Evaluation in L. loa-Endemic Areas. Clin Infect Dis. 2019;69(9):1628–30. doi: 10.1093/cid/ciz172. PubMed PMID: 30861060; PubMed Central PMCID: PMCPMC6792118.

2. Taylor HR, Munoz B, Keyvan-Larijani E, Greene BM. Reliability of detection of microfilariae in skin snips in the diagnosis of onchocerciasis. Am J Trop Med Hyg. 1989;41(4):467–71. doi: 10.4269/ajtmh.1989.41.467. PubMed PMID: 2802024.

3. Guidelines for stopping mass drug administration and verifying elimination of human onchocerciasis - Criteria and procedures, (1 January 2016, 2016).

4. Group NTDMCO. The World Health Organization 2030 goals for onchocerciasis: Insights and perspectives from mathematical modelling: NTD Modelling Consortium Onchocerciasis Group. Gates Open Res. 2019;3:1545. Epub 20190926. doi: 10.12688/gatesopenres.13067.1. PubMed PMID: 31723729; PubMed Central PMCID: PMCPMC6820451.

5. Lobos E, Weiss N, Karam M, Taylor HR, Ottesen EA, Nutman TB. An immunogenic Onchocerca volvulus antigen: a specific and early marker of infection. Science. 1991;251(5001):1603-5. doi: 10.1126/science.2011741. PubMed PMID: 2011741.

6. Gbakima AA, Nutman TB, Bradley JE, McReynolds LA, Winget MD, Hong Y, et al. Immunoglobulin G subclass responses of children during infection with Onchocerca volvulus. Clin Diagn Lab Immunol. 1996;3(1):98–104. doi: 10.1128/cdli.3.1.98-104.1996. PubMed PMID: 8770512; PubMed Central PMCID: PMCPMC170255.

7. Weil GJ, Steel C, Liftis F, Li BW, Mearns G, Lobos E, et al. A rapid-format antibody card test for diagnosis of onchocerciasis. J Infect Dis. 2000;182(6):1796–9. Epub 20001026. doi: 10.1086/317629. PubMed PMID: 11069258.

8. Golden A, Steel C, Yokobe L, Jackson E, Barney R, Kubofcik J, et al. Extended result reading window in lateral flow tests detecting exposure to Onchocerca volvulus: a new technology to improve epidemiological surveillance tools. PLoS One. 2013;8(7):e69231. Epub 20130723. doi: 10.1371/journal.pone.0069231. PubMed PMID: 23935960; PubMed Central PMCID: PMCPMC3720650.

9. Katholi CR, Toe L, Merriweather A, Unnasch TR. Determining the prevalence of Onchocerca volvulus infection in vector populations by polymerase chain reaction screening of pools of black flies. J Infect Dis. 1995;172(5):1414–7. doi: 10.1093/infdis/172.5.1414. PubMed PMID: 7594692.

10. Unnasch TR, Meredith SE. The use of degenerate primers in conjunction with strain and species oligonucleotides to classify Onchocerca volvulus. Methods Mol Biol. 1996;50:293–303. doi: 10.1385/0-89603-323-6:293. PubMed PMID: 8751366.

11. Zimmerman PA, Guderian RH, Aruajo E, Elson L, Phadke P, Kubofcik J, et al. Polymerase chain reaction-based diagnosis of Onchocerca volvulus infection: improved detection of patients with onchocerciasis. J Infect Dis. 1994;169(3):686–9. doi: 10.1093/infdis/169.3.686. PubMed PMID: 8158053.

12. Alhassan A, Makepeace BL, LaCourse EJ, Osei-Atweneboana MY, Carlow CK. A simple isothermal DNA amplification method to screen black flies for Onchocerca volvulus infection. PLoS One. 2014;9(10):e108927. Epub 20141009. doi: 10.1371/journal.pone.0108927. PubMed PMID: 25299656; PubMed Central PMCID: PMCPMC4191976.

13. Wilson NO, Badara Ly A, Cama VA, Cantey PT, Cohn D, Diawara L, et al. Evaluation of Lymphatic Filariasis and Onchocerciasis in Three Senegalese Districts Treated for Onchocerciasis with Ivermectin. PLoS Negl Trop Dis. 2016;10(12):e0005198. Epub 20161207. doi: 10.1371/journal.pntd.0005198. PubMed PMID: 27926918; PubMed Central PMCID: PMCPMC5142766.

14. Khowawisetsut L, Sarasombath PT, Thammapalo S, Loymek S, Korbarsa T, Nochote H, et al. Therapeutic trial of doxycyclin plus ivermectin for the treatment of Brugia malayi naturally infected cats. Vet Parasitol. 2017;245:42–7. Epub 20170818. doi: 10.1016/j.vetpar.2017.08.009. PubMed PMID: 28969836.

15. Bennuru S, Kodua F, Drame PM, Dahlstrom E, Nutman TB. A novel, bioinformatically informed highly sensitive NAAT assay for the diagnosis of loiasis and its use for detection of circulating cell free DNA. J Infect Dis. 2023. Epub 20230527. doi: 10.1093/infdis/jiad186. PubMed PMID: 37243712.

16. Macfarlane CL, Quek S, Pionnier N, Turner JD, Wanji S, Wagstaff SC, et al. The insufficiency of circulating miRNA and DNA as diagnostic tools or as biomarkers of treatment efficacy for Onchocerca volvulus. Sci Rep. 2020;10(1):6672. Epub 20200421. doi: 10.1038/s41598-020-63249-4. PubMed PMID: 32317658; PubMed Central PMCID: PMCPMC7174290.

17. Tritten L, Burkman E, Moorhead A, Satti M, Geary J, Mackenzie C, et al. Detection of circulating parasite-derived microRNAs in filarial infections. PLoS Negl Trop Dis. 2014;8(7):e2971. Epub 20140717. doi: 10.1371/journal.pntd.0002971. PubMed PMID: 25033073; PubMed Central PMCID: PMCPMC4102413.

18. O’Connell EM, Harrison S, Dahlstrom E, Nash T, Nutman TB. A Novel, Highly Sensitive Quantitative Polymerase Chain Reaction Assay for the Diagnosis of Subarachnoid and Ventricular Neurocysticercosis and for Assessing Responses to Treatment. Clin Infect Dis. 2020;70(9):1875–81. doi: 10.1093/cid/ciz541. PubMed PMID: 31232448; PubMed Central PMCID: PMCPMC7156770.

19. Pilotte N, Maasch J, Easton AV, Dahlstrom E, Nutman TB, Williams SA. Targeting a highly repeated germline DNA sequence for improved real-time PCR-based detection of Ascaris infection in human stool. PLoS Negl Trop Dis. 2019;13(7):e0007593. Epub 20190722. doi: 10.1371/journal.pntd.0007593. PubMed PMID: 31329586; PubMed Central PMCID: PMCPMC6675119.

20. Sears WJ, Qvarnstrom Y, Dahlstrom E, Snook K, Kaluna L, Balaz V, et al. AcanR3990 qPCR: A Novel, Highly Sensitive, Bioinformatically-Informed Assay to Detect Angiostrongylus cantonensis Infections. Clin Infect Dis. 2021;73(7):e1594-e600. doi: 10.1093/cid/ciaa1791. PubMed PMID: 33252651; PubMed Central PMCID: PMCPMC8492198.

21. Pilotte N, Thomas T, Zulch MF, Sirois AR, Minetti C, Reimer LJ, et al. Targeting a highly repetitive genomic sequence for sensitive and specific molecular detection of the filarial parasite Mansonella perstans from human blood and mosquitoes. PLoS Negl Trop Dis. 2022;16(12):e0010615. Epub 20221229. doi: 10.1371/journal.pntd.0010615. PubMed PMID: 36580452; PubMed Central PMCID: PMCPMC9833530.

22. Steel C, Lujan-Trangay A, Gonzalez-Peralta C, Zea-Flores G, Nutman TB. Immunologic responses to repeated ivermectin treatment in patients with onchocerciasis. J Infect Dis. 1991;164(3):581–7. doi: 10.1093/infdis/164.3.581. PubMed PMID: 1822959.

23. Francis H, Awadzi K, Ottesen EA. The Mazzotti reaction following treatment of onchocerciasis with diethylcarbamazine: clinical severity as a function of infection intensity. Am J Trop Med Hyg. 1985;34(3):529–36. doi: 10.4269/ajtmh.1985.34.529. PubMed PMID: 4003668.

24. Novak P, Neumann P, Pech J, Steinhaisl J, Macas J. RepeatExplorer: a Galaxy-based web server for genome-wide characterization of eukaryotic repetitive elements from next-generation sequence reads. Bioinformatics. 2013;29(6):792–3. Epub 20130201. doi: 10.1093/bioinformatics/btt054. PubMed PMID: 23376349.

25. Novak P, Neumann P, Macas J. Graph-based clustering and characterization of repetitive sequences in next-generation sequencing data. BMC Bioinformatics. 2010;11:378. Epub 20100715. doi: 10.1186/1471-2105-11-378. PubMed PMID: 20633259; PubMed Central PMCID: PMCPMC2912890.

26. Gan W, Gu Y, Han J, Li CX, Sun J, Liu P. Chitosan-Modified Filter Paper for Nucleic Acid Extraction and “in Situ PCR” on a Thermoplastic Microchip. Anal Chem. 2017;89(6):3568–75. Epub 20170308. doi: 10.1021/acs.analchem.6b04882. PubMed PMID: 28230980.

27. Rosenbohm JM, Robson JM, Singh R, Lee R, Zhang JY, Klapperich CM, et al. Rapid electrostatic DNA enrichment for sensitive detection of Trichomonas vaginalis in clinical urinary samples. Anal Methods. 2020;12(8):1085–93. Epub 20200205. doi: 10.1039/c9ay02478f. PubMed PMID: 35154421; PubMed Central PMCID: PMCPMC8837197.

28. Deer DM, Lampel KA, Gonzalez-Escalona N. A versatile internal control for use as DNA in real-time PCR and as RNA in real-time reverse transcription PCR assays. Lett Appl Microbiol. 2010;50(4):366–72. Epub 20100122. doi: 10.1111/j.1472-765X.2010.02804.x. PubMed PMID: 20149084.

29. Fischer P, Buttner DW, Bamuhiiga J, Williams SA. Detection of the filarial parasite Mansonella streptocerca in skin biopsies by a nested polymerase chain reaction-based assay. Am J Trop Med Hyg. 1998;58(6):816–20. doi: 10.4269/ajtmh.1998.58.816. PubMed PMID: 9660471.

30. Zoure HGM, Wanji S, Noma M, Amazigo UV, Diggle PJ, Tekle AH, et al. The Geographic Distribution of Loa loa in Africa: Results of Large-Scale Implementation of the Rapid Assessment Procedure for Loiasis (RAPLOA). Plos Neglect Trop D. 2011;5(6). doi: ARTN e1210 10.1371/journal.pntd.0001210. PubMed PMID: WOS:000292139600045.

31. Hendy A, Kruger A, Pfarr K, De Witte J, Kibweja A, Mwingira U, et al. The blackfly vectors and transmission of Onchocerca volvulus in Mahenge, south eastern Tanzania. Acta Trop. 2018;181:50–9. Epub 20180202. doi: 10.1016/j.actatropica.2018.01.009. PubMed PMID: 29410302.

32. Arko-Boham B, Aryee NA, Blay RM, Owusu EDA, Tagoe EA, Doris Shackie ES, et al. Circulating cell-free DNA integrity as a diagnostic and prognostic marker for breast and prostate cancers. Cancer Genet. 2019;235–236:65-71. Epub 20190423. doi: 10.1016/j.cancergen.2019.04.062. PubMed PMID: 31105051.

33. Edwards RL, Menteer J, Lestz RM, Baxter-Lowe LA. Cell-free DNA as a solid-organ transplant biomarker: technologies and approaches. Biomark Med. 2022;16(5):401–15. Epub 20220223. doi: 10.2217/bmm-2021-0968. PubMed PMID: 35195028.

34. Padilla-Martinez F, Wojciechowska G, Szczerbinski L, Kretowski A. Circulating Nucleic Acid-Based Biomarkers of Type 2 Diabetes. Int J Mol Sci. 2021;23(1). Epub 20211228. doi: 10.3390/ijms23010295. PubMed PMID: 35008723; PubMed Central PMCID: PMCPMC8745431.

35. Buttner G, Zea-Flores G, Poltera AA, Buttner DW. Fine structure of microfilariae in the skin of onchocerciasis patients after exposure to amocarzine. Trop Med Parasitol. 1991;42(3):314–8. PubMed PMID: 1801159.

36. Darge K, Lucius R, Monson MH, Behrendsen J, Buttner DW. Immunohistological and electron microscopic studies of microfilariae in skin and lymph nodes from onchocerciasis patients after ivermectin treatment. Trop Med Parasitol. 1991;42(4):361–7. PubMed PMID: 1796234.

37. Volik S, Alcaide M, Morin RD, Collins C. Cell-free DNA (cfDNA): Clinical Significance and Utility in Cancer Shaped By Emerging Technologies. Mol Cancer Res. 2016;14(10):898–908. Epub 20160715. doi: 10.1158/1541-7786.MCR-16-0044. PubMed PMID: 27422709.

38. Azad AA, Volik SV, Wyatt AW, Haegert A, Le Bihan S, Bell RH, et al. Androgen Receptor Gene Aberrations in Circulating Cell-Free DNA: Biomarkers of Therapeutic Resistance in Castration-Resistant Prostate Cancer. Clin Cancer Res. 2015;21(10):2315–24. Epub 20150223. doi: 10.1158/1078-0432.CCR-14-2666. PubMed PMID: 25712683.

39. Augustus E, Van Casteren K, Sorber L, van Dam P, Roeyen G, Peeters M, et al. The art of obtaining a high yield of cell-free DNA from urine. PLoS One. 2020;15(4):e0231058. Epub 20200406. doi: 10.1371/journal.pone.0231058. PubMed PMID: 32251424; PubMed Central PMCID: PMCPMC7135229.

